# The Effect of Sensory Reweighting on Postural Control and Cortical Activity in Parkinson’s Disease

**DOI:** 10.1101/2024.01.26.24301687

**Authors:** Maryam Sadeghi, Thomas Bristow, Sodiq Fakorede, Ke Liao, Jacqueline A. Palmer, Kelly E. Lyons, Rajesh Pahwa, Chun-Kai Huang, Abiodun Akinwuntan, Hannes Devos

## Abstract

**Aims:** Balance requires the cortical control of visual, somatosensory, and vestibular inputs. The aim of this cross-sectional study was to compare the contributions of each of these systems on postural control and cortical activity using a sensory reweighting approach between participants with Parkinson’s disease (PD) and controls.

**Methods:** Ten participants with PD (age: 72 ± 9; 3 women; Hoehn & Yahr: 2 [1.5 – 2.50]) and 11 controls (age: 70 ± 3; 4 women) completed a sensory organization test in virtual reality (VR-SOT) while cortical activity was being recorded using electroencephalography (EEG). Conditions 1 to 3 were completed on a stable platform; conditions 4 to 6 on a foam. Conditions 1 and 4 were done with eyes open; conditions 2 and 5 in a darkened VR environment; and conditions 3 and 6 in a moving VR environment. Linear mixed models were used to evaluate changes in center of pressure (COP) displacement and EEG alpha and theta/beta ratio power between the two groups across the postural control conditions. Condition 1 was used as reference in all analyses.

**Results:** Participants with PD showed greater COP displacement than controls in the anteroposterior (AP) direction when relying on vestibular input (condition 5; p<0.0001). The mediolateral (ML) COP sway was greater in PD than in controls when relying on the somatosensory (condition 2; p = 0.03), visual (condition 4; p = 0.002), and vestibular (condition 5; p < 0.0001) systems. Participants with PD exhibited greater alpha power compared to controls when relying on visual input (condition 2; p = 0.003) and greater theta/beta ratio power when relying on somatosensory input (condition 4; p = 0.001).

**Conclusions:** PD affects reweighting of postural control, exemplified by greater COP displacement and increased cortical activity. Further research is needed to establish the temporal dynamics between cortical activity and COP displacement.

## Introduction

Postural imbalance is a hallmark symptom of Parkinson’s disease (PD)^1^. Several clinical tests, such as the Berg Balance Scale, the Tinetti Balance Test, the BESTest, or the Mini-BESTest, have been developed and validated to identify balance impairments and determine the risk of falls in individuals with PD^2^. Although these tests include assessments of both static and dynamic postural control^3^, they provide limited insights into the specific sensory systems that contribute to postural control in PD. The Sensory Organization Test (SOT) was designed to quantitatively evaluate the ability to maintain postural control by incorporating visual, vestibular, and somatosensory inputs^4^. The SOT uses sensory reweighting in which the brain adjusts the relative importance of different sensory systems to sustain balance^5,6^. This sensory reweighting is compromised in PD, indicating difficulties in distinguishing and selecting reliable information from various sensory systems to ensure postural control^4,7^. Particularly, individuals with PD show increased postural sway when relying predominantly on the inputs of the somatosensory systems and vestibular systems^8–11^. These studies suggest that individuals with PD exhibit impaired central processing of somatosensory and vestibular information, resulting in increased reliance on vision to maintain balance. Yet, the cortical processes involved with sensory reweighting of postural control in PD have not been investigated.

Mobile neuroimaging technology is increasingly used to elucidate the cortical processes related to postural control in healthy adults and adults with neurological conditions ^12,13,14^. Electroencephalography (EEG) utilizes electrodes placed on the scalp to measure voltage potential differences between two locations on the scalp^15^. The neuronal oscillations captured using EEG can be divided into delta (0 – 4 Hz), theta (4 – 7 Hz), alpha (8 – 12 Hz), beta (13 – 30 Hz), and gamma (>30 Hz) frequency bands. Within these bands, alpha power and the ratio of slow (theta) to fast wave (beta) activity are of particular interest to study the cortical processes of postural control^16,17^. Changes in occipital alpha power appear to reflect the sensory and movement-related information processing of postural control^18^. In neurotypical young adults, occipital alpha power increased when standing with the eyes closed^19,20^. Theta/beta ratio power reflects cognitive activity related to attention and working memory processes^21,22^. Theta/beta ratio power increased while standing with the eyes closed and while standing while performing a dual task in neurotypical young adults^20^. In PD, participants with balance impairments showed decreased mid-frontal and mid-cerebellar theta power while standing with the eyes open compared to those with no balance problems and neurotypical older adults^23^. The results of this previous study^23^ warrant further assessment to elucidate the cortical processes involved in the sensory organization of postural control in PD.

The aim of this study was to compare the effect of sensory reweighting on postural control and cortical activity between participants with PD and older adults. We hypothesized that participants with PD will exhibit greater CoP displacement and increased cortical activity in the SOT compared to older adults.

## Methods

### Participants

Ten participants with a diagnosis of idiopathic PD were recruited from the Parkinson’s disease and Movement Disorder Center. Eleven control participants were age- and sex-matched with PD participants and were recruited from the community. Inclusion criteria were the ability to comprehend and follow instructions in English, ability to stand without assistive devices, scoring more than 20 on the Montreal Cognitive Assessment (MoCA)^24^, and the participant with PD being in the medication on state. Exclusion criteria were a diagnosis of severe cognitive impairment or dementiahi, visual acuity or loss of visual fields that cannot be resolved with corrective lenses, severe head and trunk dystonia or dyskinesia in the medication on state, blepharospasm, unpredictable motor fluctuations, deep brain stimulation, and presence of any musculoskeletal conditions which can affect standing and balance activities. This cross-sectional study was approved by Institutional Review Board (#00148555).

### Protocol

All participants provided informed consent before starting the study procedures. Relevant demographic information such as age, sex, education level, disease duration, disease severity (measured using the Hoehn and Yahr (H&Y) scale)^25^ and medication details were extracted from medical records. Medication dose was converted into a Levodopa Equivalent Daily Dose (LEDD)^26^. The Montreal Cognitive Assessment (MoCA) was administered as a general screen of cognitive function^24^. In addition, we recorded the participant-reported number of falls in the six months prior to their visit. All experiments were conducted when participants were in their optimal medication state (ON), about 45 minutes after medication intake.

We used the virtual reality-based Comprehensive Balance Assessment and Training (VR-COMBAT) system to administer the virtual reality SOT (VR-SOT)^27^. The VR-COMBAT system includes a processing computer (Alienware, Dell), a VR-integrated head-mounted device (VR-HMD) from HTC VIVE Pro Eye, and VR tracking sensors (Steam VR Base Stations, HTC). The HTC VIVE Pro Eye features dual OLED displays (2,880 × 1,600 pixels) with a 90 Hz refresh rate and a 110-degree field of view. The Steam VR software (version 1.13, Valve) links the computer and VR headset. The head-mounted device integrates with an AMTI Optima force plate (Watertown, MA), synchronized for precise measurements during trials.

### Experimental Procedure

Participants were instructed to remove their shoes and socks and stand barefoot on an AMTI Optima force plate (Watertown, MA, United States). They were guided to position their heel centers 17.5 cm apart with their feet oriented at 14°. Each participant was fitted with a safety harness attached to an overhead anchor for security during testing. A trained spotter was positioned behind each participant to ensure safety throughout the procedure.

The VR-SOT comprises six different conditions that mimic the six conditions of the Equitest® SOT. Condition 1 serves as a benchmark for assessing static balance on a stable surface with fixed VR surroundings. The panels in the VR environment do not move. Participants can use the feedback of the visual, somatosensory, and vestibular systems to keep their balance. Condition 2 provides the same balance testing condition as to condition 1. However, participants must rely on somatosensory inputs to remain upright since the VR surroundings are blackened out^28^. In condition 3, the surrounding panels are moving in the anteroposterior direction with a maximum of 20 degrees and a maximum velocity of 15 degrees/sec. This condition creates a conflict between normal input from the somatosensory and vestibular systems and the visual information from the moving VR panels. Conditions 4, 5, and 6 mirror the parameters of conditions 1, 2, and 3, respectively. However, in conditions 4, 5, and 6, a foam (19 x 15 x 1.5 inch) is placed between the feet and the force plate, thus challenging the somatosensory system. In condition 4, participants must rely on visual inputs to maintain balance, whereas in condition 5, participants must rely on vestibular inputs^28,29^. Condition 6 generates a conflict between visual, somatosensory and vestibular systems.

Each of the six conditions comprised three trials, with each trial lasting 20 seconds. Between each trial, a 5-second break was given. Data of the three trials were averaged.

### Data Capture and Processing

The center of pressure (COP) data were obtained from the force plate at 200 Hz and processed using the MATLAB software (MathWorks, Natick, MA)^30^ to compute the following parameters^31^:

♣ Mean displacement in the anterior-posterior (AP) or medial-lateral (ML) direction (MeanAP or MeanML): The average displacement of the COP from its mean position in the AP/ML direction.
♣ Mean velocity in the AP or ML direction (VelAP or VelML): The average speed at which the COP moves in the AP/ML direction.
♣ Average frequency in the AP or ML direction (MfAP or MfML): The rotational frequency (in Hz) of the COP as it completes a full circle with a radius equal to the mean displacement.

### EEG Data Acquisition and Processing

To capture cortical activity, we used the mobile EXG system from Mentalab (Munich, Germany). Eight electrodes (seven dry brush electrodes on the scalp and one flat wet reference electrode) were wired to the Mentalab Explore hub attached to the back of the EEG net. The Explore device captures the scalp electrical activity at 500 Hz and transmits the signals to the laptop via Bluetooth. Electrode placement included the midline channels Fz, Pz, Cz, Oz, the frontal channels Fpz and Fp1, the central channel Fcz, and the reference electrode on the right mastoid TP10, according to the International 10-20 system. Impedance was kept below 50 kΩ for the recording. Feasibility testing ensured that placing the VR apparatus over the EEG net did not compromise EEG recording.

The EEG data underwent filtering from 0.1 to 30 Hz using the EEGlab^32^ plugin within the MATLAB software^33^. To reduce noise, EEG recordings were trimmed 10 seconds before the trials began and 10 seconds after the last trial ended. The continuous EEG data were segmented into six datasets for each condition. EEGLAB^32^ automatic channel rejection was used to initially detect the noisy channels. In detail, the pop_rejchan() function from EEGLAB was used, in which the probability of each EEG channel was calculated as the rejection measurement with z-score threshold as 5. Then, the data were visually inspected to mark those channels with extreme noise. Finally, the Artifact Subspace Reconstruction (ASR) method^34^ from the EEGLAB clean_rawdata plugin (https://github.com/sccn/clean_rawdata) was used to reject bad data periods. The standard deviation cutoff for removal of bursts was chosen from 1 to 20 (mean/standard deviation: 8.61/6.15). The cleaned data were manually inspected again to be included into power calculation. A dataset was excluded from further data analysis if any of its channels was rejected as bad channel. To calculate the theta (4–7 Hz), alpha (8–13 Hz), and beta (13–30 Hz) frequency band power for all six conditions, EEGlab’s Spectopo function was used to extract power spectral density from all electrodes, which uses Welch’s method on the 1 s epochs with 50% overlap between its calculation windows.

### Statistical Analysis

All statistical analyses were conducted using SAS software (version 9.4, Cary, NC). Normality of data distributions was assessed through the Shapiro–Wilk test and visual inspection of the histogram and Q-Q plots. We used Fisher’s Exact tests and independent t-tests to compare demographic and clinical variables between groups.

We calculated differences in sensory reweighting on COP and cortical activity between participants with PD and healthy controls (HC) using linear mixed models. We used a random intercept term and a subject-specific coefficient to adjust for correlation between measures within subjects. The linear mixed model included the main effects of group (PD – HC), condition (1 – 6), and the interaction effect of group*condition. To minimize the risk of type 1 errors, we only compared (1) condition 2 to condition 1 to evaluate the reweighting of postural control to the somatosensory system; (2) condition 4 to condition 1 to evaluate the reweighting of postural control to the visual system; (3) and condition 5 to condition 1 to evaluate the reweighting of postural control to the vestibular system^29,35^. We only report the results of the interaction effect. Finally, we correlated COP with EEG measures for each group separately using Pearson r correlations. The significance threshold for all analyses was set at a = 0.05.

## RESULTS

### Demographic and Clinical Variables

No differences were found in demographic and clinical characteristics between groups, except for the MoCA score (Table 1). Participants with PD scored slightly lower on the MoCA compared to controls. The disease duration and the H & Y stages indicated that participants with PD were in the very mild to moderate stage of the disease.

**Table 1:**
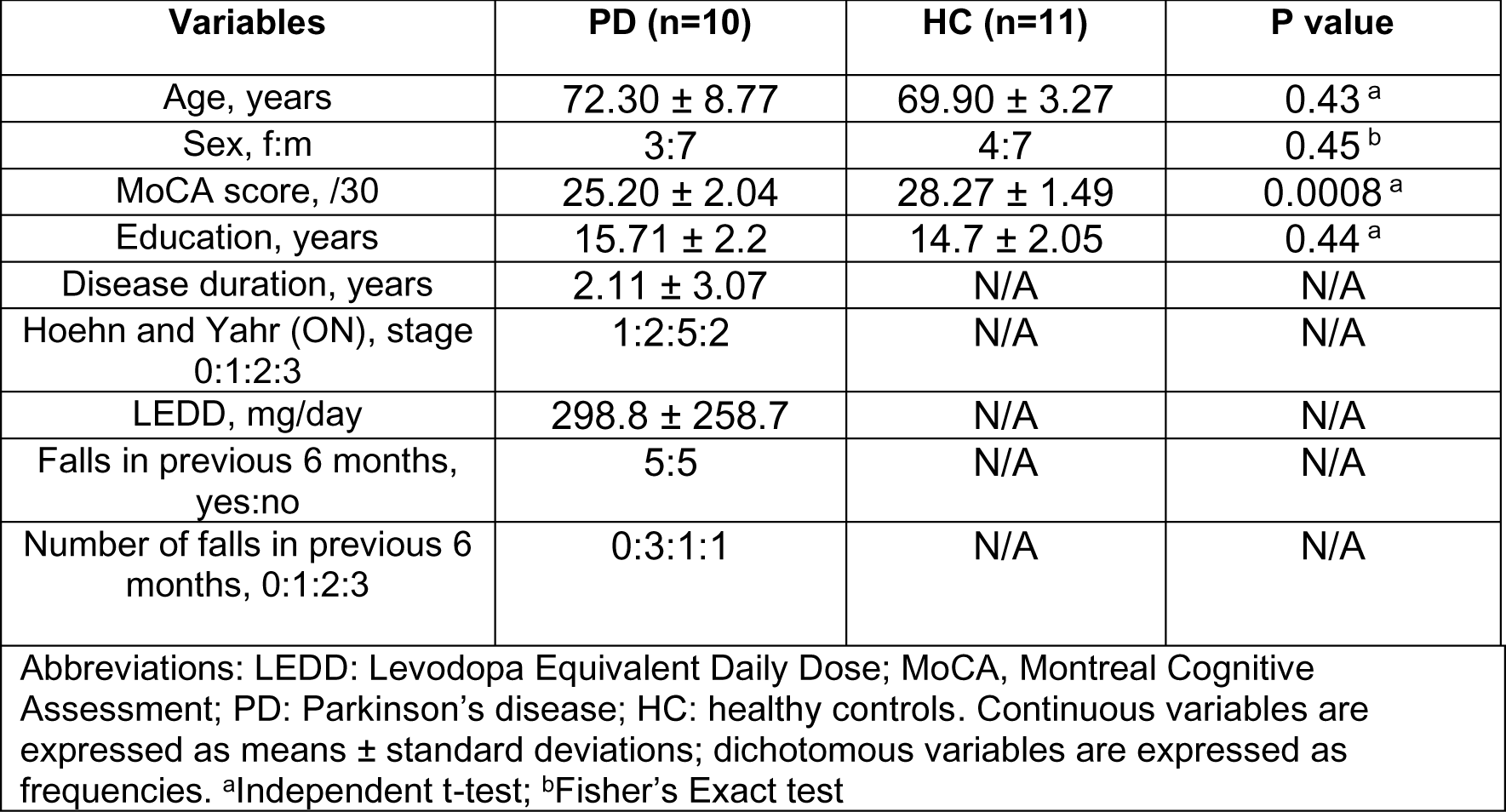
Comparison of demographic and clinical characteristics between groups.

**Table 2:**
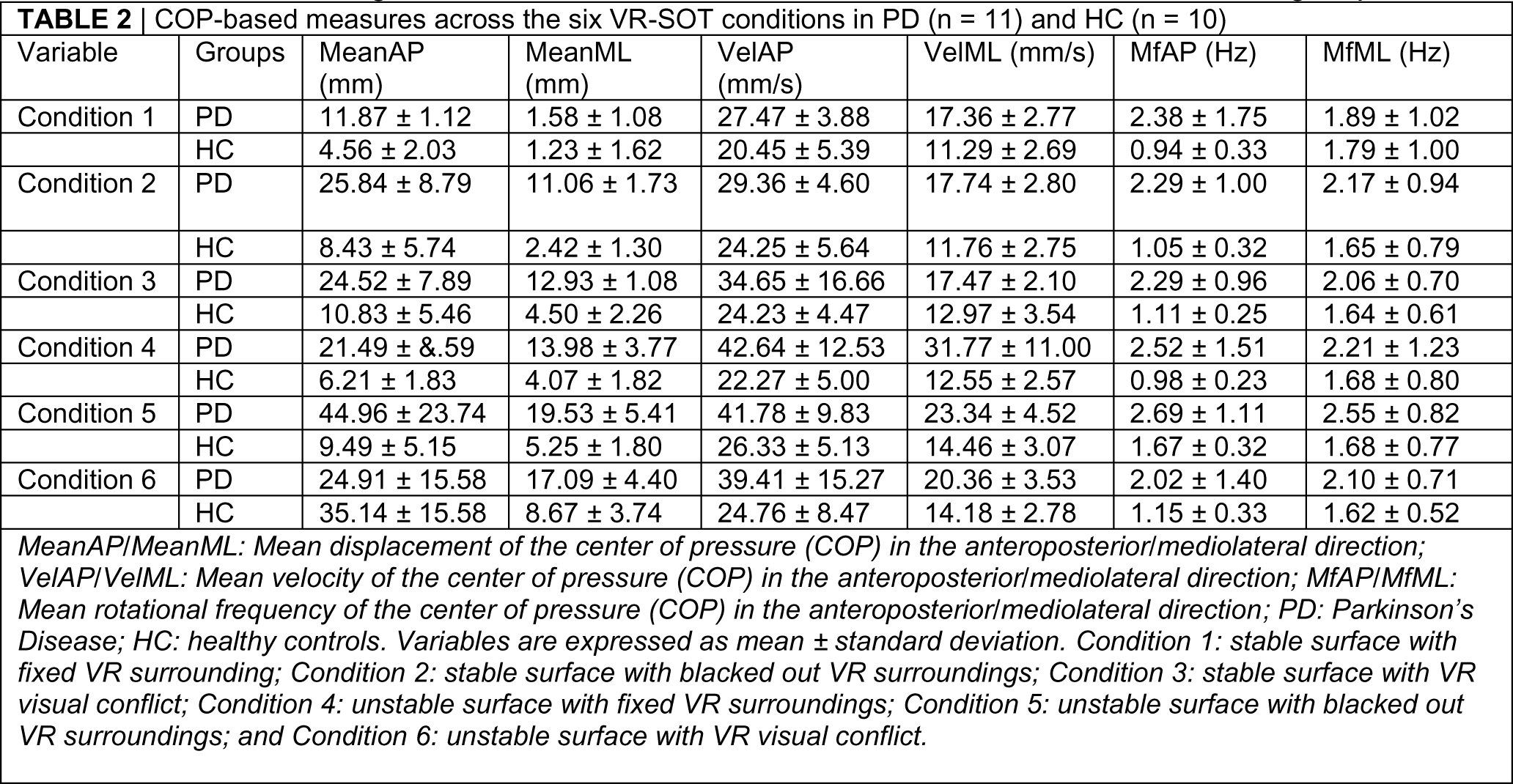
Shows the average COP variables across the six conditions in the PD and HC groups.

During the postural balance conditions, two participants with PD and one control lost their balance and requested support from the spotter. One person with PD discontinued in condition 5; one control discontinued in condition 4. Another person with PD lost balance during condition 4 but completed all conditions.

### COP variables

#### COP displacement

The analysis of mean AP displacement demonstrated a significant group by condition interaction effect (F = 8.31, p < 0.001). Post-hoc analysis revealed that this interaction was significant for condition 5 (p < 0.001) relative to condition 1. Participants with PD exhibited greater displacement in the AP direction compared to controls when relying on vestibular inputs to maintain balance (Figure 1A).

**Figure 1:**
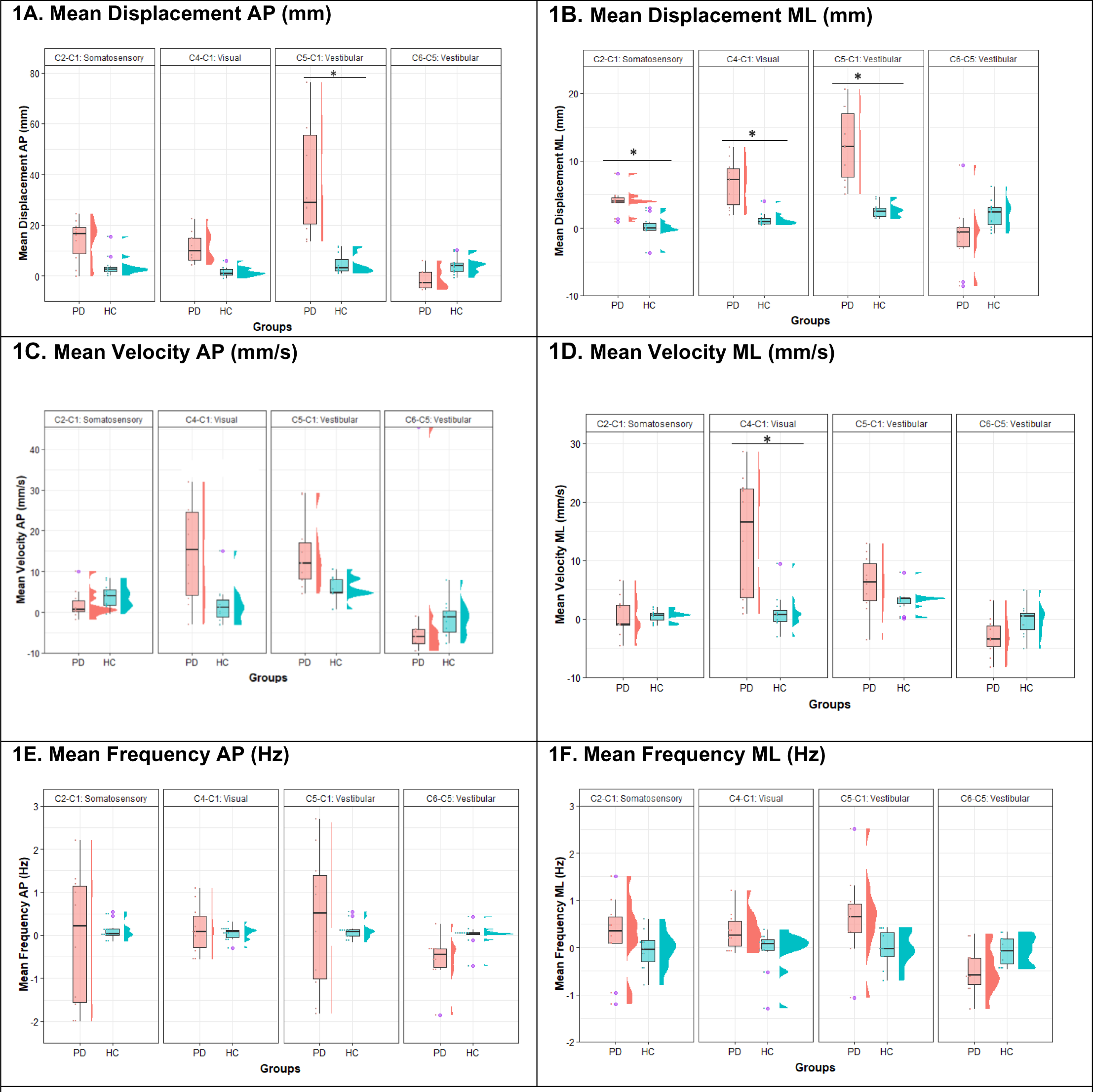
Comparison of center of pressure data using box-and-whisker and raincloud plots between participants with PD (n = 10) and neurotypical controls (n=11). AP, anteroposterior; ML, mediolateral. Abbreviations: PD: Parkinson’s disease; HC: healthy controls.

Similarly, the analysis of mean ML displacement demonstrated a significant group by condition interaction effect (F = 7.25, p < 0.0001). Post-hoc analysis indicated that this interaction was significant for condition 2 (p = 0.03), condition 4 (p = 0.002), and condition 5 (p < 0.0001) relative to condition 1. Individuals with PD displayed greater ML displacement compared to controls when relying on either somatosensory, visual, or vestibular inputs (Figure 1B).

### COP velocity

The analysis of AP velocity approached significance for group by condition interaction effect (F = 2.01, p = 0.08). No post-hoc effects were therefore calculated.

The analysis of ML velocity demonstrated a significant group by condition interaction effect (F = 7.64, p < 0.001). Post-hoc analysis indicated that this interaction was significant for condition 4 (p < 0.001) relative to condition 1. Individuals with PD displayed greater velocity of the COP in the ML direction compared to controls when the relative contribution of the visual system to postural control was being tested (Figure 1D).

### COP frequency

The analysis of frequency outcomes did not demonstrate any significant group by condition interaction effects (Figure 1E and Figure 1F).

### EEG variables Alpha Power

The analysis of alpha power demonstrated a significant group by condition interaction effect (F = 3.50, p = 0.005). Post-hoc analysis revealed that this interaction was specifically significant for condition 4 (p = 0.003) relative to condition 1. Participants with PD (5.97 ± 7.23 µV/Hz^2^) exhibited increased alpha power compared to controls (3.49 ± 4.41 µV/Hz^2^) when relying on visual inputs of postural control (Figure 2A).

**Figure 2:**
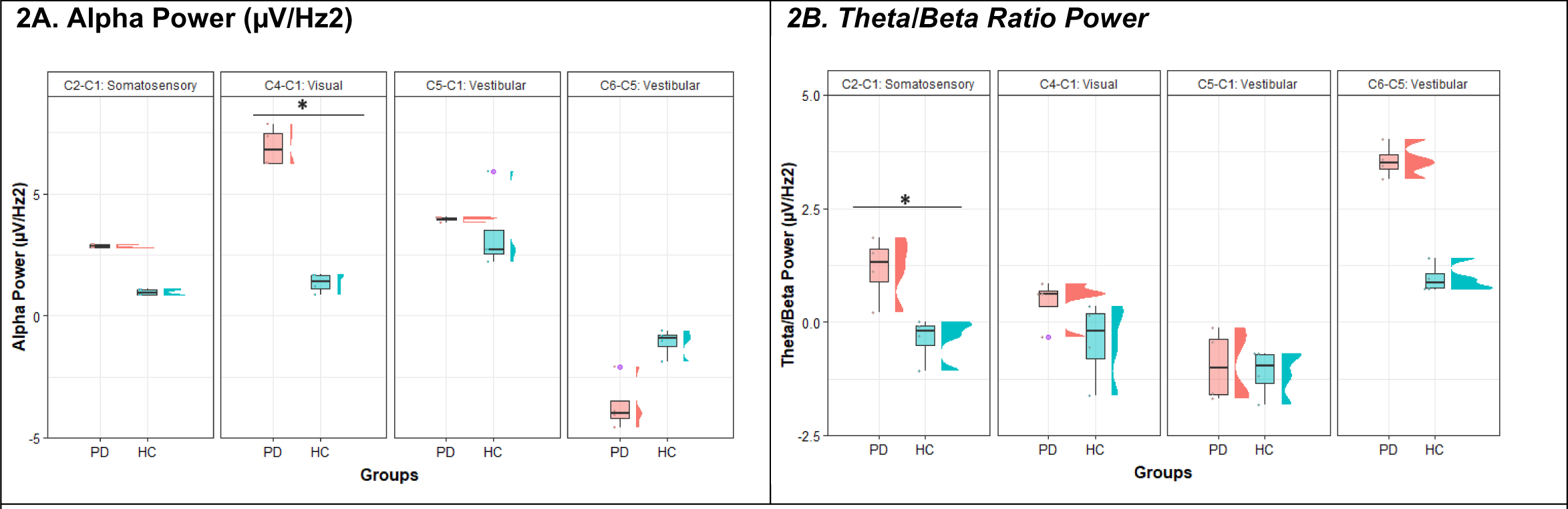
Comparison of EEG power using box-and-whisker and raincloud plots between participants with PD (n = 10) and healthy controls (n=11). Abbreviations: PD: Parkinson’s disease; HC: healthy controls.

Since participants with PD exhibited cognitive impairment, we repeated the linear mixed model while adjusting for MOCA scores. Although MOCA scores were associated with alpha power (F = 4.59, p < 0.001), the interaction effect of group by condition remained significant (F = 3.74; p = 0.0007), with post-hoc effects for condition 4 (p = 0.0004) relative to condition 1.

### Theta/Beta Ratio Power

Similarly, the analysis of theta/beta ratio power demonstrated a significant group by condition interaction (F = 3.77, p = 0.003). Post-hoc analysis indicated that this interaction was significant for condition 2 (p = 0.01) relative to condition 1. Individuals with PD (5.14 ± 3.27) displayed increased theta/beta ratio power compared to controls (2.95 ± 2.51) when the relative contribution of the somatosensory system was being tested.

We repeated the linear mixed models while adjusting for MOCA scores. MOCA scores were not associated with theta/beta ratio power (F = 0.59; p = 0.79), and the interaction effect of group by condition remained significant (F = 3.20; p = 0.005), with post-hoc effects for condition 2 (p = 0.03) relative to condition 1.

Figure 3 shows the topographical maps of alpha power when the visual contribution to postural control (condition 4 – condition 1) and theta/beta ratio power when the somatosensory contribution was being tested (condition 2 – condition 1) in participants with PD and controls.

**Figure 3:**
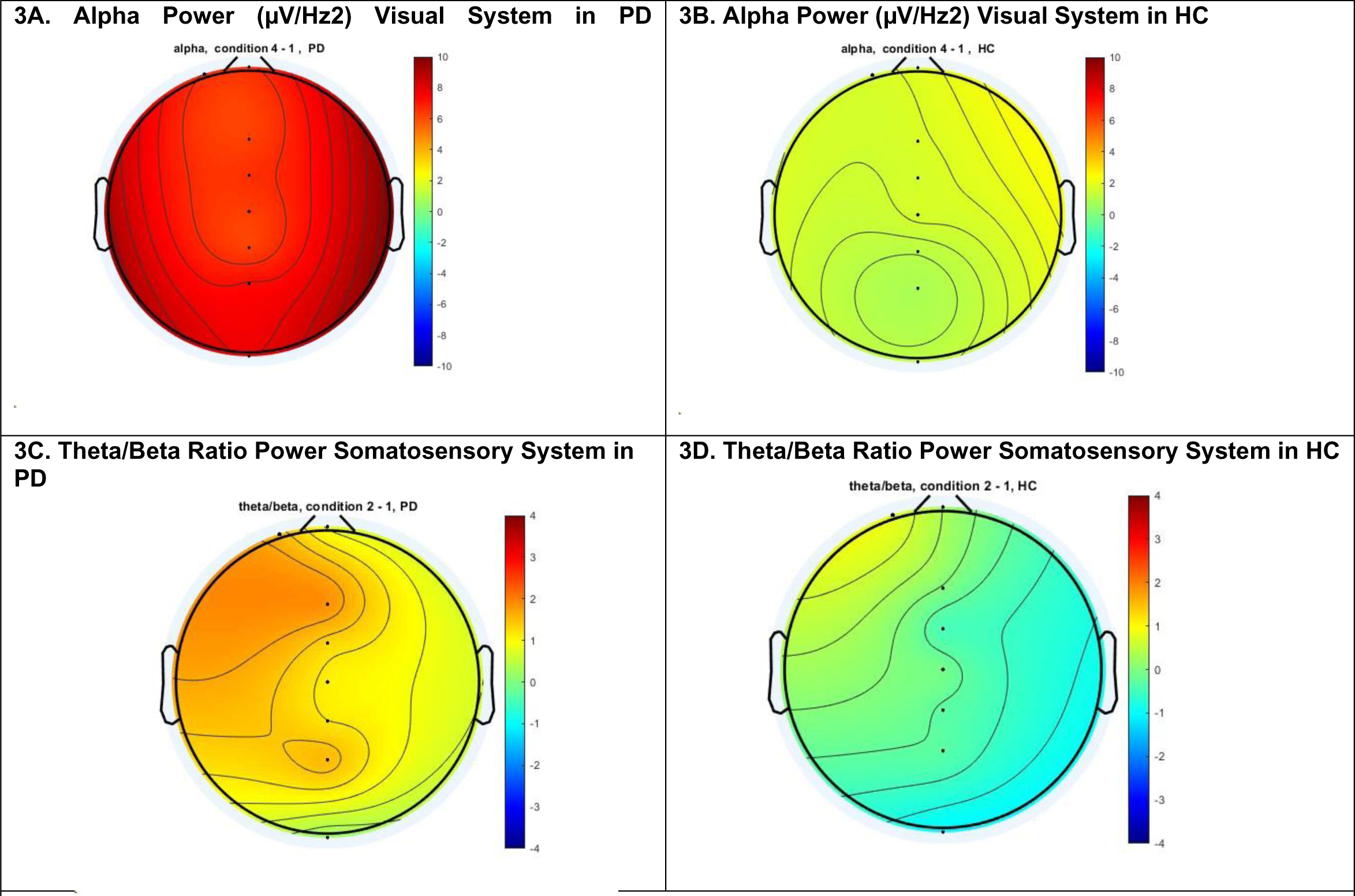
Topographical maps of alpha power (µV/Hz2) during visual system testing and theta/beta ratio power during somatosensory system testing in the PD group (n = 10) and ***the HC group (n = 11).*** Abbreviations: PD: Parkinson’s disease; HC: healthy controls.

## Discussion

The purpose of this study was to examine the impact of sensory reweighting of postural control on center of pressure (COP) and cortical activity in individuals with Parkinson’s disease (PD) compared to healthy older adults. We observed two key findings: (1) participants with PD displayed greater ML displacement when relying on somatosensory, visual, or vestibular inputs; and (2) participants with PD exhibit greater cortical activity when relying on somatosensory and visual inputs of postural control compared to healthy controls.

Participants with PD exhibited greater COP displacement and velocity particularly in the ML direction, compared to the control group. While increased postural sway in the anteroposterior (AP) direction is more noticeable with higher postural control task difficulty, both PD and control groups responded similarly to sensory adjustments in the AP direction. Previous studies have indicated that individuals in the early stage of PD demonstrated the ability to use sensory inputs for postural control in the AP direction effectively^36–38^. However, they showed reduced ability to quickly adapt postural control to changing sensory conditions^39^. In contrast, individuals with PD showed increased COP displacement in the ML direction when relying on either somatosensory, visual, or vestibular inputs. Previous studies suggesting that ML sway is more sensitive than AP sway in detecting postural instability, detecting disease progression, or risk of falls in PD^40,41^. Our results extend to these findings by delineating sensory-specific postural impairments in PD.

We hypothesized that the increased COP displacement during sensory reweighting is linked to increased cortical activity. Indeed, participants with PD exhibited increased alpha power when relying primarily using the visual system, and increased theta/beta ratio power when primarily relying on the somatosensory system for postural control. The increased alpha power may reflect either impaired central processing of visual cues, or difficulties resolving visual cues from conflicting somatosensory cues. Previous research has shown increased alpha power when the inputs from the visual and somatosensory systems are incongruent in healthy adults^19^.

Theta/beta ratio power increased in participants with PD when relying on somatosensory cues^42^. Occlusion of the dominant visual system likely prompted participants with PD to focus more attention on maintaining postural control, resulting in increased theta/beta ratio power^42^. Even after adjusting for MOCA scores in the statistical models, participants with PD continued to display increased cortical activity. This implies that factors beyond cognitive impairment may contribute to the observed cortical activity changes in PD. In line with the Compensation-Related Utilization of Neural Circuits Hypothesis (CRUNCH) model^43^, individuals with PD might engage additional cortical regions or increase activation in specific areas to compensate for postural control impairments.

While the study presents the first link between cortical activity and sensory reweighting in PD, caution is necessary due to the relatively small sample size and the VR-SOT used in this study ^27^. Many VR systems currently on the market (e.g., Bertec®, Virtualis®, UprightVR) or used for research have found the VR-SOT to be reliable compared to traditional SOT^44,45^. Future studies should encompass larger sample sizes, a wider range of PD severities, and subtypes for a more comprehensive understanding how PD affects cortical processes of sensory reweighting. Particularly, the robustness of our findings should be tested in a group of PD patients with no cognitive impairments. The study results are also sufficiently encouraging to explore the use of EEG as an early marker of balance impairment in PD. Future studies should evaluate the temporal association between changes in cortical activity and balance impairment in PD.

### Conclusion

In conclusion, current study sheds light on the impact of PD on the sensory organization of postural control. Participants with PD showed difficulties adjusting postural control to the relative inputs of the visual, somatosensory, and vestibular systems, particularly in the mediolateral direction. Participants with PD exhibit increased cortical activity when relying on somatosensory and visual inputs to maintain postural control. Future studies should investigate the usefulness of electroencephalography as markers of balance impairments in PD.

## Data availability statement

Authors will make available all raw data supporting their conclusions without undue delay.

## Ethics statement

The research involving human participants underwent a review and received approval from the Institutional Review Board (IRB) at the. All participants provided their explicit written consent to take part in this study.

